# Higher energy requirement during weight-loss maintenance on a low- versus high-carbohydrate diet: secondary analyses from a randomized controlled feeding study

**DOI:** 10.1101/19001248

**Authors:** Cara B Ebbeling, Lisa Bielak, Paul R Lakin, Gloria L Klein, Julia MW Wong, Patricia K Luoto, William W Wong, David S Ludwig

## Abstract

**Background:** Longer-term feeding studies suggest that a low-carbohydrate diet increases energy expenditure, consistent with the carbohydrate-insulin model of obesity. However, the validity of methodology utilized in these studies, involving doubly-labeled water, has been questioned.

**Objective:** The aim of this study was to determine whether dietary energy requirement for weight-loss maintenance is higher on a low-versus high-carbohydrate diet.

**Methods:** The study reports secondary outcomes and exploratory analyses from a feeding study in which the primary outcome was total energy expenditure. After attaining a mean Run-in weight loss of 10.5%, 164 adults with pre-weight-loss BMI of ≥25 were randomly assigned to Test diets containing Low (20%), Moderate (40%) or High (60%) carbohydrate for 20 weeks. Calorie content of Test diets was adjusted to maintain individual body weight within 2 kg of the post-weight-loss value. In analyses by Intention-to-Treat (ITT, study completers, n=148) and Per Protocol (PP, those achieving the weight-loss maintenance target, n=110), we compared estimated energy requirement from 10 to 20 weeks on the Test diets using ANCOVA. Insulin secretion was assessed pre-weight-loss as insulin concentration 30 minutes following 75 grams oral glucose (Insulin-30).

**Results:** Estimated energy requirement was higher in the Low *vs* High group by models involving ITT (ranging from 181 [CI 8–353] to 223 [40–406] kcal/d; *P*≤0.04) and PP (ranging from 245 [43–446] to 295 [91–499] kcal/d; *P*≤0.02). This difference remained significant in sensitivity analyses accounting for change in adiposity and possible non-adherence. In observational analyses, pre-weight loss Insulin-30 predicted adverse change in body composition following weight loss.

**Conclusions:** Energy requirement was higher on a low-versus high-carbohydrate diet during weight-loss maintenance, commensurate with total energy expenditure. These data are consistent with the carbohydrate-insulin model and lend qualified support for the validity of the doubly-labeled water method with diets varying in macronutrient composition.

## INTRODUCTION

The independent effect of dietary composition on energy expenditure remains a topic of controversy. According to the carbohydrate-insulin model (CIM) of obesity, the high ratio of blood insulin-to-glucagon concentration in the postprandial period with consumption of a high glycemic load diet partitions metabolic fuels toward fat storage (1, 2). As a result, hunger may increase and (under some conditions, such as post-weight loss) energy expenditure may decrease relative to a low glycemic load diet. Because reduced energy expenditure following weight loss may predispose to weight regain (3-5), research into the dietary determinants of metabolic rate holds both scientific and clinical significance.

A recent meta-analysis reported little effect of dietary carbohydrate-to-fat ratio on energy expenditure (6), but the included studies had a median duration of < 1 week. As previously reviewed (2), the adaptation to a low-carbohydrate diet takes at least 2 to 3 weeks, limiting inferences about chronic macronutrient effects that can be drawn from these very short trials. The few prior studies of at least 2.5 weeks duration consistently showed a numerical advantage favoring the low-carbohydrate diet (2), but each of these had important methodological limitations, such as low statistical power, lack of randomization and physical confinement (*e.g.*, in respiratory chambers) confounding activity-related energy expenditure.

In the longest feeding study addressing this question (7), we reported that total energy expenditure (TEE) was about 200 to 250 kcal/d higher on a low-*vs* high-carbohydrate Test diet throughout 20 weeks of weight-loss maintenance, as determined using doubly-labeled water (DLW) methodology. However, the validity of DLW with diets varying in macronutrient composition has recently been called into question (8).

The aim of the present study was to examine estimated energy requirement (EER) for maintenance of stable weight following weight loss in our study, based on the energy provided to participants in carefully controlled Test diets. If TEE increases with reduction in dietary carbohydrate and DLW methodology is valid for measuring TEE when comparing different macronutrient diets, we would expect to see corresponding dietary effects on EER.

## METHODS

### Overview of parent study design and original findings

This study presents secondary and exploratory (*post hoc*) analyses from a feeding trial for which the methods (trial design, participants, dietary interventions, sample size, randomization), participant flow, adverse events and primary outcome were previously reported (7, 9, 10). Briefly, 164 participants with overweight or obesity who lost at least 10% of their body weight during the Run-in phase on a hypocaloric diet were randomly assigned to low- (LOW, 20% carbohydrate, 60% fat), moderate- (MOD; 40% carbohydrate, 40% fat) or high- (HIGH, 60% carbohydrate, 20% fat) carbohydrate Test diets controlled for protein (20%). During the 20-week Test phase, dietary energy provided to participants in prepared meals was adjusted with the aim of keeping weight within 2 kg of the post-weight loss, pre-randomization baseline value. TEE was measured using DLW at four time points: 1) pre-weight loss (PRE), 2) start of trial (START, weeks −2 to 0, post-weight-loss), 3) midpoint of Test phase (MID, weeks 8 to 10) and 4) end of Test phase (END, weeks 18 to 20). The primary finding of the trial was that TEE was significantly greater on LOW *vs* HIGH in an Intention-to-Treat model (ITT: 209 kcal/d, n=162, *P*=0.002 for overall group effect) and a Per Protocol model that excluded participants who did not achieve weight stability at 10 or 20 weeks (PP: 278 kcal/d, n=120, overall *P*<0.001).

We previously conducted a preliminary analysis in the PP group (n=120, including 10 participants who achieved weight stability at MID but did not complete the trial), comparing change in estimated energy intake from START to the average of MID (10 weeks) and END (20 weeks) using dietary data for the days when we assessed TEE (7). Change in energy intake increased in a pattern consistent with the dietary effect on TEE, though without significant group differences (HIGH 139, MOD 175, LOW 269 kcal/d, overall *P*=0.36). This pattern strengthened as expected among those in the highest tertile of insulin secretion at PRE (37, −24, 340 kcal/d, respectively, overall *P*=0.05). However, as discussed in our initial report, these preliminary analyses were imprecise and inaccurate, with probable bias against those with higher energy requirements, thereby limiting scientific inference.

### Conceptual approach for current analyses

For the current study, we considered four potential reasons for imprecision and inaccuracy in the initial estimates of energy requirements used to calculate energy intake: 1) excessive variability in the START estimate used in models of change; 2) the limited time frame (using dietary data only from the days when we assessed TEE) for evaluating energy intake during the Test phase; 3) unaddressed factors affecting EER, including provision of additional snacks to some individuals to assist with weight-loss maintenance and 4) change in body composition affecting energy balance calculations.

To begin our exploration of these issues, we extracted data from food production sheets throughout the Test phase on daily dietary energy provided to every participant, as periodically adjusted to maintain weight loss within the target range. Visual inspection revealed large changes (>500 kcal/d) in dietary energy provided for many participants in all 3 diet groups from the weight stabilization period at the end of the Run-in phase through the first few weeks of the Test phase, demonstrating that our initial estimates of energy requirements were imprecise. By MID (week 10 of the Test phase), estimates of energy requirements had stabilized, with relatively few participants requiring substantial adjustments in dietary energy to maintain weight loss from that point through END (week 20). This imprecision would not have biased the primary study outcome involving TEE, because the initial dietary energy level for each participant was established prior to randomization and there was no significant difference in body weight among the diet groups during the assessment periods. However, imprecision in the START value for energy intake would erode power for the change models originally reported (11, 12).

As an alternative approach to the inherent limitations of an imprecise baseline (START value) in measurement of a change variable, we examined EER with general linear models (ANCOVA) adjusted for baseline covariates that would plausibly influence energy requirements (such as age, sex and weight). We focused on the daily average energy provided from 10 to 20 weeks as our most accurate measure of EER, with primary interest in the HIGH *vs* LOW diet comparison in the PP analysis to maximize power. Consistent with the approach used with our original TEE outcome, we calculated EER per kg and normalized the results as kcal/d using the average START weight of our participants (82 kg). We examined diet group differences in EER at START and during weeks 10 through 20 of the Test phase, with and without adjustment for the START value. In sensitivity analyses, we explored how changes in body fat mass might influence EER and how possible non-adherence to energy prescription might influence EER.

### Assessment of Test diet energy

Details regarding the dietary interventions were published previously (9, 10). In brief, the hypocaloric Run-in diet comprised 45% of energy from carbohydrate, 35% from fat, and 25% from protein. Eucaloric Test diets were controlled for protein and varied in carbohydrate-to-fat ratio as indicated above. Standardized menus were calculated for 2000-kcal Run-in and Test diets using Food Processor Nutrition Analysis Software (ESHA Research Inc., Salem, OR) with energy distributed across breakfast (450 kcal), lunch (650 kcal), dinner (650 kcal), and an evening snack (250 kcal). Data for each menu item were exported from the ESHA Food Processor to Excel (Microsoft, Redmond, WA), and gram weights were imported from Excel into SAS (SAS Institute Inc., Cary, NC). In SAS, 2000-kcal menus were scaled to coincide with individualized energy levels, and food production sheets (1 sheet per participant per meal or snack) were generated to specify gram portions of each menu item.

#### Estimating and adjusting Run-in and Test diet energy levels

Individualized energy levels were estimated and then adjusted when necessary, but not more frequently than every two weeks. To inform adjustments, body weight was measured daily using Wi-Fi scales (Withings Inc., Cambridge, MA) synced with a study-specific online portal (SetPoint Health, Needham, MA). At the beginning of the Run-in phase (PRE), energy levels were set at 60% of estimated needs (13), and then adjusted to achieve targeted weight loss. Energy levels for weight stabilization at the end of the Run-in phase were estimated based on rate of weight loss over 20 days: energy intake during weight loss (kcal/day) + rate of weight loss (kg/day × 7700 kcal/kg). During the Test phase, energy levels were adjusted when deviation from the START anchor weight exceeded ±2 kg and/or the slope of weight regressed on time was ≥15 g per day over 14 days.

Some participants received unit bars (100 kcal per bar with diet-specific carbohydrate-to-fat ratio) and/or *ad libitum* snacks, in addition to the meals and snacks listed on food production sheets. The purpose of providing unit bars was to: 1) replace some of the meal calories, when large portions were a barrier to consuming all provided food and 2) immediately adjust energy levels, before meal adjustments could be implemented according to established production cycles, to achieve weight-loss maintenance (±2 kg of START anchor weight). The purpose of providing foods for *ad libitum* snacks (n=11) was to halt continued weight loss in participants who were already consuming large meals. Examples of snack foods (for each diet) included: banana, skim milk (HIGH); bagel chips, chocolate chips, apple, banana, nut butters (MOD); nuts, nut butters, dark chocolate, whole milk (LOW). We conservatively estimated energy content of *ad libitum* snacks at 200 kcal per day, for the days when participants (n=11) received snacks. In preliminary analyses, we noted similar study outcomes with energy content estimated at 500 kcal per day (data not presented).

#### Quantifying unconsumed energy

Data on food consumption recorded in the online portal were used to calculate daily unconsumed energy, which totaled < 5% throughout the study. For supervised meals, unconsumed menu items were weighed, and gram amounts were entered into the portal by food service staff. Menu data exported from the ESHA Food Processor to Excel were used to create a “food library,” interfaced with the portal, for converting gram amounts to kcal. For unsupervised take-out meals, percentages of menu items consumed were recorded by participants, using a form in the online portal that was prepopulated with daily menus from food production sheets, so that unconsumed energy could be calculated as follows: energy provided – (energy provided × percentage consumed). Unconsumed energy during supervised and unsupervised meals was summed to obtain a total for each day. Food consumption data for calculating unconsumed energy were not available electronically for cohort 1 (n=25 in ITT, n=18 in PP), prior to developing the online portal, and assumed to be 0 (this methodological limitation is addressed in a sensitivity analysis).

#### Calculating EER

An EER for each participant was calculated as the average daily energy level during weeks 10 through 20 of the Test phase. The calculation included energy in weighed meals and snacks (as specified on food production sheets), *ad libitum* snacks (200 kcal/d), and unit bars (based on the number provided), with correction for unconsumed energy in weighed meals and snacks. The first 10 weeks of the Test phase was considered adequate time for physiological adaptations to the Test diets, that could affect energy metabolism and fluctuations in body weight (2), and fine-tuning initially imprecise estimates of energy levels for weight-loss maintenance. We also calculated EER for the first day of the Test phase (EER at START), to obtain insight regarding the level of imprecision and inaccuracy in the initial estimates of energy requirements, and for use as a baseline covariate in a statistical model.

### Assessment of body composition

We assessed body composition by dual-energy X-ray absorptiometry (DXA, Discovery A, Hologic Inc., Bedford, MA, USA) and isotope dilution. Data from DXA, the more precise method, were available for PRE, START and END. Data from isotope dilution were available for the same time points and also MID, allowing assessment of adiposity from weeks 10 through 20 of the Test phase which was the exact timeframe of interest for determining EER (after the initial 10-week period of physiological adaptation). Total body water was estimated using the isotope dilution space for ^18^O (calculated as previously described) (9), divided by 1.01 (to correct for binding to non-exchangeable sites) (14). Total body water was divided by 0.73 to estimate fat-free mass (FFM). Fat mass (FM) was calculated by subtracting FFM from total body weight. Percent body fat was calculated as: FM / body weight × 100%.

### Statistical analyses

For all summary and inferential computations, we used SAS 9.4 (SAS Institute, Cary, NC).

#### Descriptive data

We inspected raw distributions of EER during the Test phase for the ITT and PP groups and compared raw distributions and descriptive data (mean and median) with those of TEE.

#### Variability in Estimated Energy Requirement at START

We used partial correlation analysis to determine whether excessive variability in EER might have obscured the effect of diet on change in EER in our preliminary analyses (7). Controlling for diet group, we evaluated partial correlation of the residuals from models comparing EER at START with EER during the Test phase (MID through END), and TEE at START with TEE during the Test Phase (average of MID and END).

#### Diet effect on Estimated Energy Requirement

The analytic framework for statistical inference on EER and other outcomes was the general linear model (GLM) including ANCOVA. We evaluated EER at START (with body weight at START included as a covariate) and EER during the Test phase (with and without EER at START as a covariate). To be consistent with the approach in our prior study (7), the reported models include diet group assignment and a design variable (a polytomous covariate labeled cohort, which captured all combinations of study site, cohort, and enrollment wave, including 11 categories). Because inclusion of this variable utilizes 10 degrees of freedom, and we have no reason to hypothesize confounding by cohort, a model without this adjustment was evaluated. Other variables in the primary model included sex, age at randomization, weight loss during the Run-in phase (expressed as a percentage of PRE body weight), START weight, and START TEE. One participant who developed a medical condition (hypothyroidism) that affects energy expenditure was excluded in the final analysis plan on an *a priori* basis from the primary outcome in our prior study (7, 9). We present models with and without exclusion of this individual. The outcome was the Test phase average of EER from weeks 10-20, modified from our original change (pre-to-post) analyses, in consideration of the rationale above, and as further addressed in **Figure 1**.

**Figure 1.**
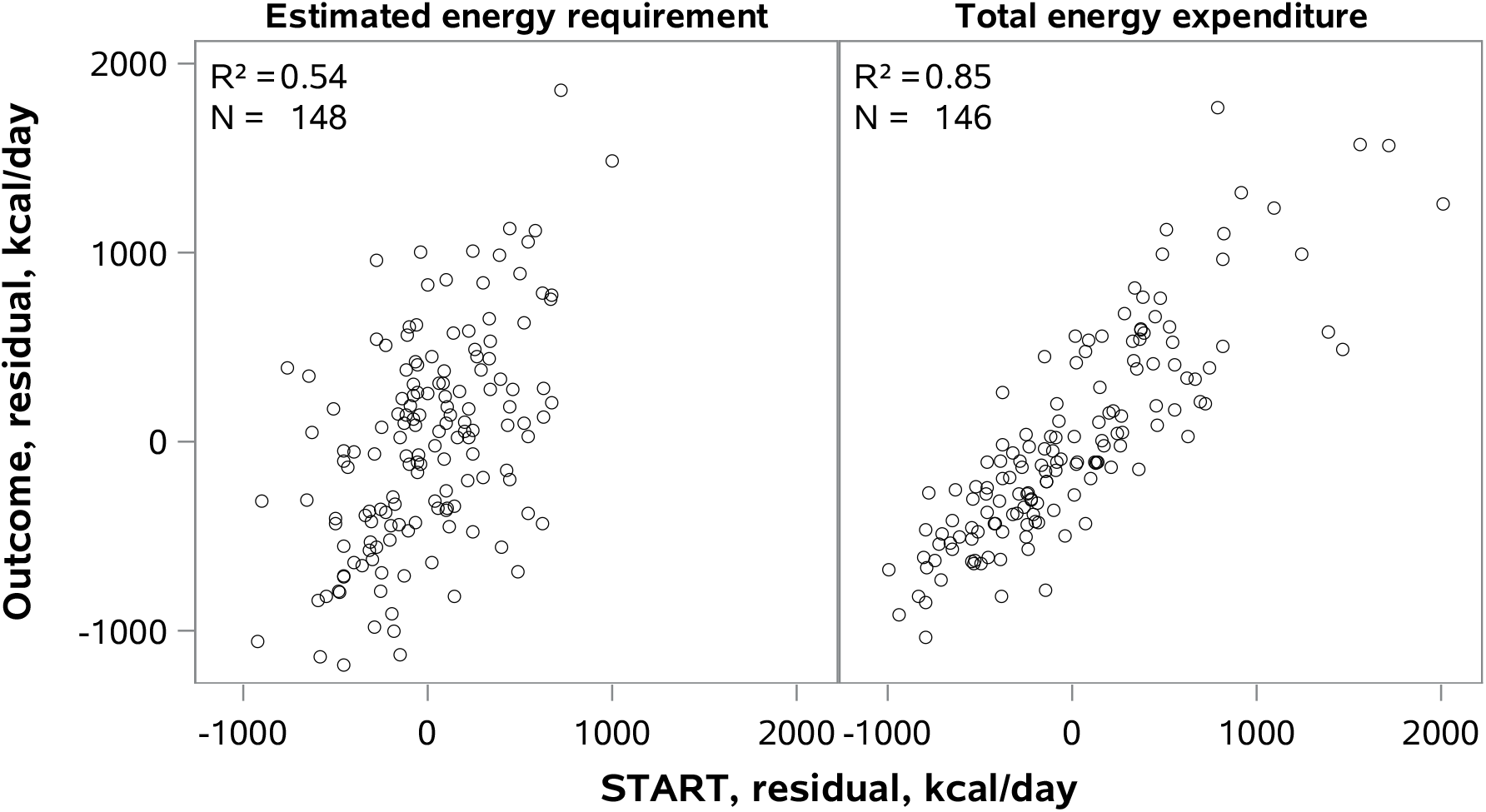
Variability in estimated energy requirement (EER) and total energy expenditure (TEE). The correlation between the baseline (START) and outcome measurements adjusted for diet was substantially lower for EER (Panel A) compared to TEE (Panel B), providing rationale for using ANCOVA rather change models for EER.

From parameters of the fitted models, taking account of all data, we tested two hypotheses. First, that the outcome was uniform across all diet groups, using an *F* test with two degrees of freedom and a *P* ≤ 0.05 as a threshold for significance. The HIGH – LOW comparison was equivalent to a test for linear trend by carbohydrate proportion, given the equal increments of carbohydrate content (60%, 40%, 20%) across Test diets. In this second test, the null hypothesis was zero difference between HIGH and LOW in a two-sided Student’s *t* test.

We conducted four sensitivity analyses using GLM (ANCOVA) to explore the potential effects of changes in body composition and non-adherence on EER during weight-loss maintenance. These analyses were based on our most conservative estimate of EER in the weight stable PP group (Table 2, model 2). For every kilogram increase or decrease in FM from START to END, assessed by DXA, we subtracted or added 55 kcal/d (7700 kcal/kg ÷ 140 days, the relevant time period). Similarly, for change in FM from MID to END, assessed by isotope dilution, we subtracted or added 110 kcal/d (7700 kcal/kg ÷ 70 days, the relevant time period). As a proxy measure of non-adherence, we defined energy discrepancy as the ratio of EER-to-TEE and excluded participants with energy discrepancy in the top quintile (those most likely to have under-consumed provided foods) and bottom quintile (those most likely to have consumed foods off protocol). In a final model, we excluded individuals in cohort 1, for whom we had no food consumption data in the online portal to calculate unconsumed energy as a measure of non-adherence.

**Table 1.** Participant characteristics.

| Characteristic | All Enrolled <sup>a</sup><br>N=164 | Intention-to-Treat <sup>b</sup><br>N=148 | Per Protocol <sup>c</sup><br>N=110 |
| --- | --- | --- | --- |
| *Categorical Variables, No (%)* |  |  |  |
| Sex |  |  |  |
| Men | 49 (29.9%) | 45 (30.4%) | 33 (30.0%) |
| Women | 115 (70.1%) | 103 (69.6%) | 77 (70.0%) |
| Hispanic Ethnicity | 25 (15.2%) | 21 (14.2%) | 18 (16.4%) |
| Race |  |  |  |
| White | 128 (78.0%) | 116 (78.4%) | 84 (76.4%) |
| Black | 17 (10.4%) | 16 (10.8%) | 11 (10.0%) |
| Asian | 5 (3.0%) | 5 (3.4%) | 4 (3.6%) |
| Unknown/Other | 14 (8.5%) | 11 (7.4%) | 11 (10.0%) |
| *Continuous Variables, mean (SD)* |  |  |  |
| Age, years | 38.04 (14.37) | 38.62 (14.42) | 39.83 (13.98) |
| Weight, kg | 91.46 (18.17) | 91.22 (18.23) | 89.49 (16.56) |
| Height, cm | 167.69 (9.99) | 167.88 (10.04) | 167.34 (10.28) |
| Body Mass Index (BMI), kg/m<sup>2</sup> | 32.37 (4.83) | 32.20 (4.77) | 31.82 (4.17) |
| Weight Loss, % of PRE body weight | 10.45 (1.68) | 10.49 (1.59) | 10.47 (1.53) |
| Total Energy Expenditure at START, kcal/d | 2661 (547) | 2651 (557) | 2663 (559) |

<sup>a</sup> Included in observational analyses of Insulin-30 and body weight and body composition
<sup>b</sup> Study completers
<sup>c</sup> Study completers who achieved weight-loss maintenance target

**Table 2.**
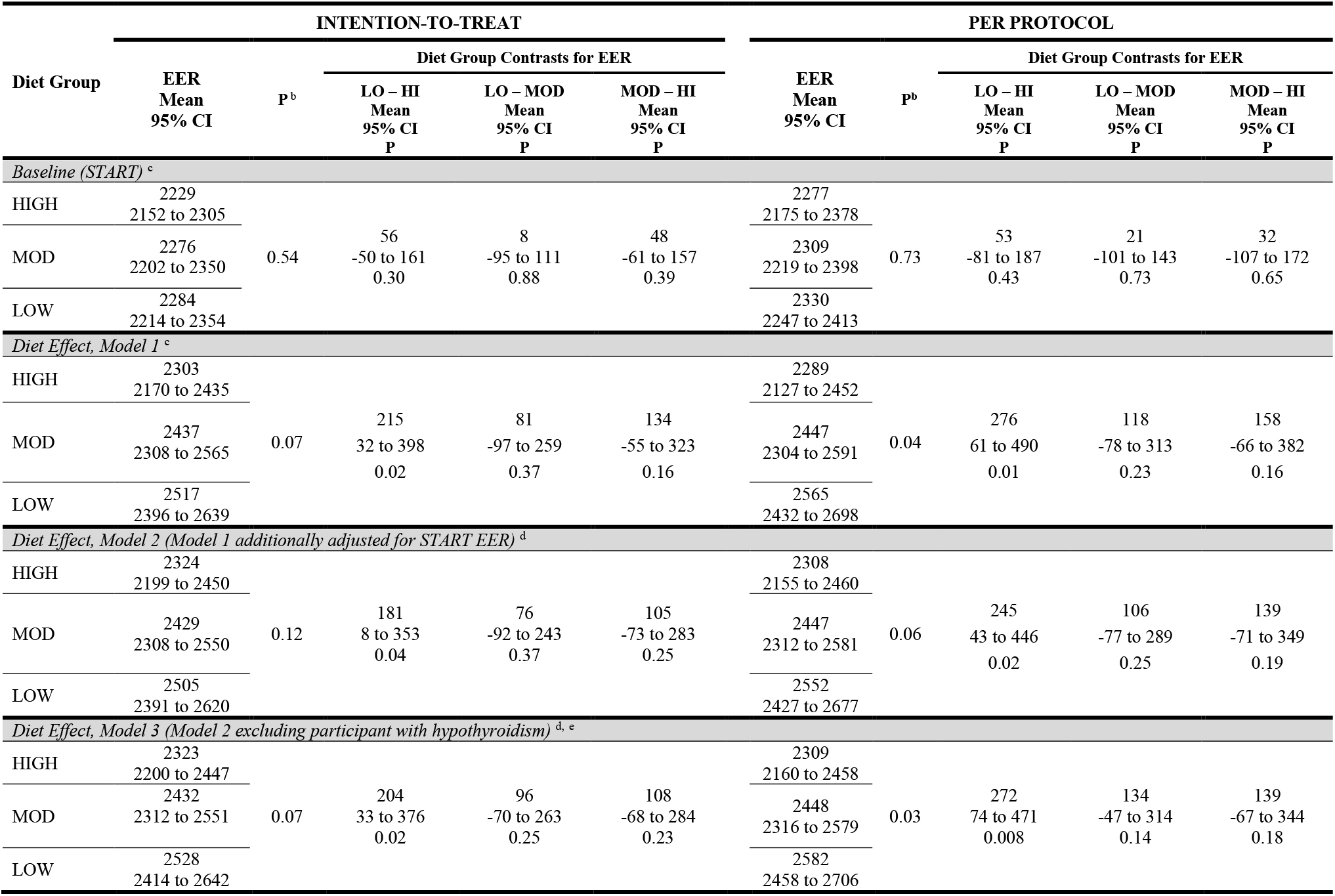
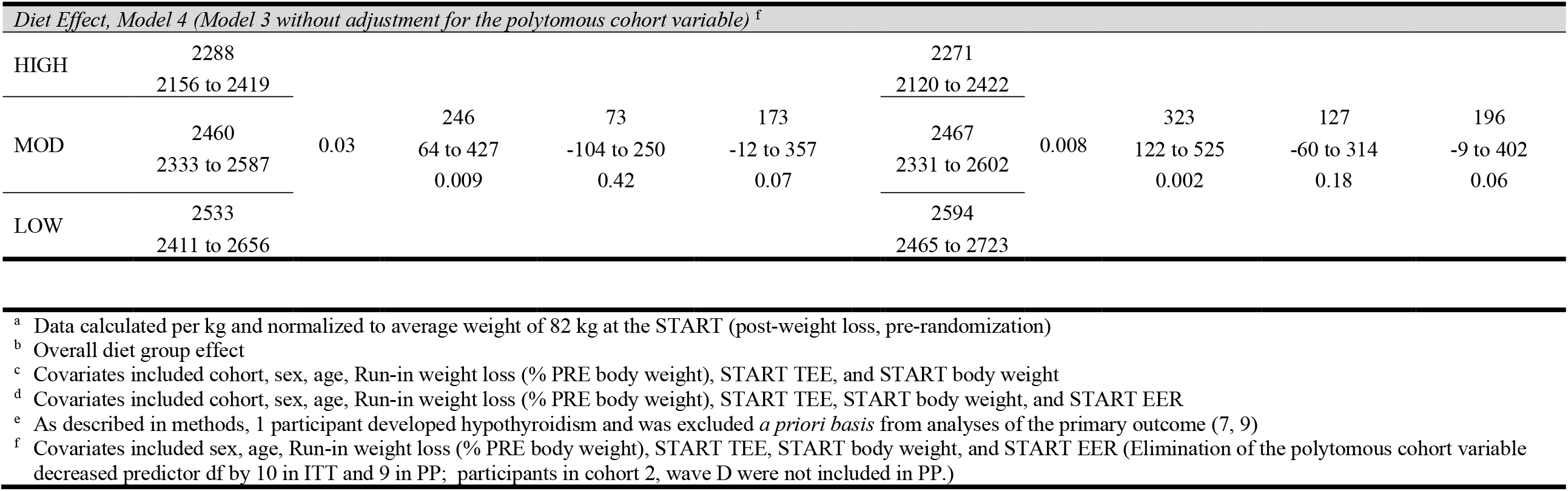
Effects of Test diets on Estimated Energy Requirement (EER) during weight-loss maintenance ^a^

#### Changes in body composition

We analyzed change in percent body fat from START to END with DXA, and also from MID to END with isotope dilution. These models included only design variables (diet group, cohort). In cross-sectional and prospective observational analyses involving the Run-in phase, we evaluated the associations of Insulin-30 (measured at PRE) with PRE body weight or percent body fat by DXA, and with change in percent body fat by DXA (from PRE to START). These models include participant characteristics (sex, age).

### Ethics

The study protocol was approved by the institutional review board at Boston Children’s Hospital and registered at ClinicalTrials.gov NCT02068885.

## RESULTS

### Descriptive data

We randomly assigned 164 participants to a diet group for the Test phase. Of these, 148 completed the trial and were included in the ITT analyses. Among the completers, 110 achieved weight-loss maintenance and were included in PP analyses. **Table 1** summarizes baseline data describing the cohort, including those who completed the weight loss Run-in phase (for analysis of Insulin-30 and body composition) and those in the ITT and PP groups (for the trial outcomes). About two-thirds of the cohort were women, mean age was ∼30 years, and mean BMI at PRE was ∼32 kg/m^2^. **Supplemental Figure 1** illustrates raw distributions of EER for the ITT and PP groups. Overall, the median and mean values of EER were 89.2% and 87.7% of TEE, respectively.

### Variability in Estimated Energy Requirement at START

To determine whether excessive variability might have obscured the effect of diet on energy requirements in our preliminary analyses of change (7), we compared EER at START with EER measured from weeks 10 through 20 of the Test phase. As shown in **Figure 1**, the partial R^2^ after adjusting for diet group (0.54) was much weaker than the partial R^2^ involving TEE (0.85). These findings suggest that analytic models of change have adequate power for evaluating TEE but not EER (11, 12), providing rationale for using an alternative approach (ANCOVA).

### Diet effect on Estimated Energy Requirement

**Table 2** shows EER by diet group in the ITT and PP analyses. At START, EER did not differ by diet. From weeks 10 through 20 of the Test phase, EER was significantly higher in LOW *vs* HIGH, ranging from a mean of 181 to 323 kcal/d in models with varying covariate structure. In sensitivity analyses (**Table 3)**, this diet effect remained robust after accounting for concurrent change in body composition, excluding individuals for whom the EER-to-TEE ratio raised the possibility of non-adherence, and additional exclusion of individuals in cohort 1 lacking non-adherence data from the online portal. The nominal order of effect by group, with MOD intermediate between LOW and HIGH, showed a pattern similar to that of TEE. The ratio of EER-to-TEE did not differ by diet group (**Supplemental Figure 2**), indicating no selective non-adherence or bias in group comparisons.

**Table 3.**
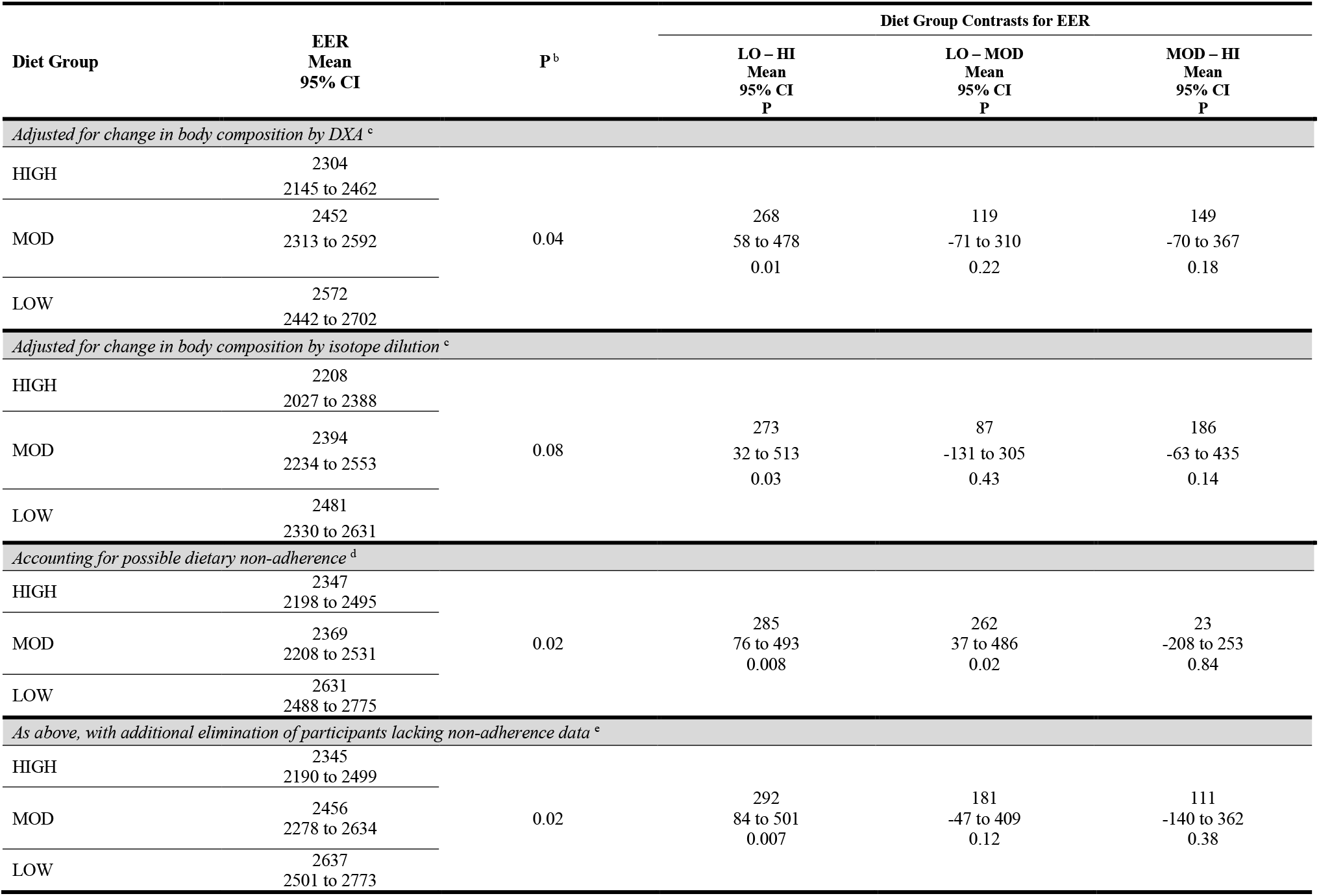
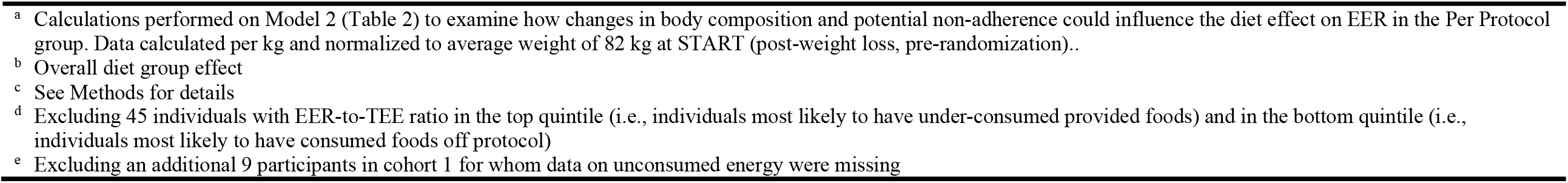
Sensitivity analysis of Estimated Energy Requirement (EER).^a^

### Changes in body composition

As shown in **Supplemental Table 1**, there were no significant diet group differences in adiposity by DXA or isotope dilution throughout the study.

In cross-sectional analyses, Insulin-30 was strongly associated with PRE body weight (4.4 kg per 100 µU/mL increase in Insulin-30, *P* for linear trend = 0.0005; **Figure 2A**) and adiposity (1.2% body fat per 100 µU/mL increase in Insulin-30, *P* for linear trend = 0.005; data not depicted). Insulin-30 also predicted change in adiposity during weight loss, with percent body fat decreasing less from PRE to START among individuals in the top versus bottom quintiles of Insulin-30 (−3.1% *vs* −3.8%, *P*=0.0085; *P* for linear trend = 0.002; **Figure 2B**). This prospective association was moderately attenuated, but remained statistically significant, with further adjustment for PRE body weight, BMI or adiposity. (However, inclusion of these adiposity measures may over-correct the models, due to potential collinearity with Insulin-30.)

**Figure 2.**
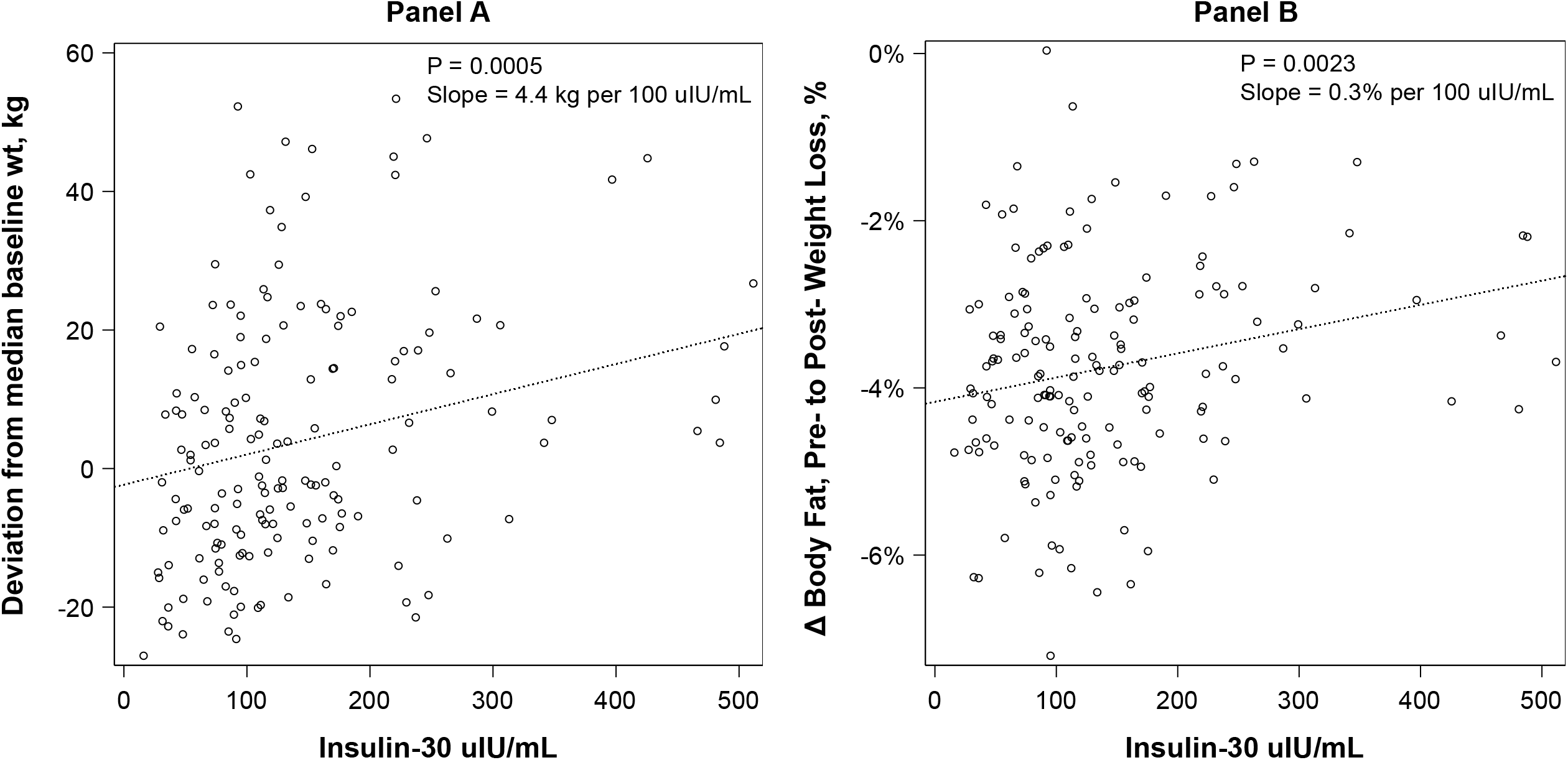
Associations of Insulin-30 with body weight and change in change in body fatness. Individuals with high Insulin-30 prior to weight loss have higher body weight in a cross-sectional analysis (Panel A) and a more adverse response to weight loss (proportionately less fat loss) in a prospective analysis (Panel B).

## DISCUSSION

In this analysis of a large feeding study, we observed higher estimated energy requirement on a low- *vs* high-carbohydrate diet during weight-loss maintenance. The magnitude of this effect (about 200 to 300 kcal/d, or ≥ 50 kcal/d for every 10% decrease in carbohydrate as a proportion of total energy) and the numerical order across groups (LOW > MOD > HIGH) are commensurate with previously reported changes in TEE (7), supporting the CIM. If reproducible and generalizable, this finding may inform the scientific understanding of how dietary composition affects metabolism and the design of more efficacious long-term obesity treatment.

These results also have relevance to outpatient metabolic study methods. In a recent analysis of an observational pilot study, Hall *et al*. (8) questioned the validity of DLW methodology to compare diets differing in carbohydrate-to-fat ratio, in part due to the “theoretical possibility that … [differential] fluxes through biosynthetic pathways” could inflate measured energy expenditure on diets with lower carbohydrate content. However, their estimates of isotopic trapping through *de novo* lipogenesis, the pathway of greatest potential concern, appear overstated, and DLW has worked well in animals with diets varying widely in macronutrient ratio, including obligate carnivores (7, 15). The congruence in dietary effect on energy intake and expenditure from our trial provide qualified validation for the use of DLW in human diet studies, though the possibility of other, unrecognized biases cannot be excluded. In contrast to the theoretical concerns involving DLW, whole room calorimetry – the other gold standard method – has been shown to underestimate adaptive thermogenesis (16) because of inherent constraints on physical activity energy expenditure (a confounding issue in the analyses of Hall *et al*. (8)). Recognizing that reduction in dietary carbohydrate has been hypothesized to attenuate adaptive thermogenesis with weight loss (2, 7), macronutrient studies utilizing whole room calorimetry may yield results biased against low-carbohydrate diets. Indeed, the prior validation study (16) found a better correspondence between dietary calorie titration and TEE – the approach we used here – for DLW *vs* whole room calorimetry under several physiological conditions.

In observational analyses, pre-weight loss insulin secretion was strongly associated with greater body weight and adiposity before weight loss, and prospectively with an adverse change in body composition (a lesser decrease in adiposity) following weight loss, potentially predisposing to weight regain. Although we cannot rule out reverse causation and confounding, the findings are consistent with other lines of investigation free from such limitations. According to the CIM, increased primary insulin secretion (versus secondary hyperinsulinemia in compensation for insulin resistance) partitions metabolic fuels away from oxidation and instead into storage, lowering energy expenditure and promoting adiposity. Indeed, individual predisposition to high insulin secretion has been linked to weight gain in translational research (17), a cohort study (18), a Mendelian randomization study (19) and several clinical trials (20, 21). These relationships appear to be strongly attenuated on a low-glycemic load diet, as was also reported for TEE among individuals with high insulin secretion in our recent trial (7). In contrast, DIETFITS found no effect modification involving insulin secretion for weight loss on lower-fat *vs* lower-carbohydrate diets, but that null finding might relate to the focus on reducing sugar and other processed carbohydrates throughout the trial, resulting in a low glycemic load in both diet groups (22). Thus, our current study provides additional data on a novel diet-phenotype interaction and highlights a high-risk subgroup that may do especially well with dietary carbohydrate restriction, similar to findings from DiOGenes and other trials involving fasting glucose or insulin resistance (23).

Strengths of this study include relatively large size and long duration for a feeding trial, demonstration of weight stability during the Test phase, state-of-the-art methods to produce nutrient-controlled diets and monitor quality control, concurrent measurement of body composition, and sufficient power to conduct informative sensitivity analyses. The main limitation is non-adherence to the Test diets. This methodological issue, common to all long-term outpatient feeding studies, could lead to an overestimation of the diet effect on energy requirement under two conditions: if individuals on the low-versus high-carbohydrate diet consumed less of the provided food than reported when not under direct observation; or if those on the high-versus low-carbohydrate diet consumed more food off protocol. Either of these scenarios might arise if the low-carbohydrate diet were less palatable or more satiating.

Conversely, the high-carbohydrate diet was substantially lower in energy density; the diet effect could be underestimated if participants in that group had difficulty consuming the larger volume of food. However, we designed the diets to be as similar as possible in types of foods included, cooking methods and palatability (10). Moreover, we saw no discrepancy in the ratio of EER-to-TEE across diet groups, nor evidence of overall bias in a sensitivity analyses excluding individuals with EER-to-TEE ratio in the highest quintile (for whom energy intake might have been overestimated) and in the lowest quintile (for whom energy intake might have been underestimated). Moreover, the findings strengthened in the PP analyses, involving participants who demonstrated successful weight-loss maintenance as an objective measure of compliance (the opposite would be expected if non-adherence contributed importantly to the observed effect).

Other study limitations include the inherent imprecision of methods for measuring small changes in body composition during weight-loss maintenance, and possible inaccuracy arising from changes in body water on diets differing in macronutrient content. However, on the latter issue, any changes in body water resulting from reduction in dietary carbohydrate would stabilize after a few weeks, allowing for an unconfounded measurement of body composition between 10 and 20 weeks of the Test phase, the relevant period for our calculations of energy requirements. Furthermore, our estimates of energy requirements vary based on covariate structure of the analytic models and other assumptions, and the 3-way diet comparison is not significant in some models. However, the comparison between the low- and high-carbohydrate diet was consistently significant as hypothesized in multiple models and sensitivity analyses. In light of the foregoing, our estimates of the magnitude of the diet effect on energy requirements should be interpreted cautiously.

Because of the inherent limitations of outpatient feeding studies discussed here, some suggest that the only informative diet studies are those conducted on metabolic wards (24), but these too have major limitations. For logistical and financial reasons, ward studies rarely exceed a few weeks in duration – too short to distinguish transient adaptive processes from the chronic metabolic effects of macronutrients (2, 25). Ward studies also entail an artificial environment, constraint on spontaneous physical activities, and the psychobiological effects of social isolation and other stresses. Moreover, even with presumably maximum control, substantial “unaccounted energy” – the basis of criticisms of our trial by Hall *et al*. (26) – may occur, as was seen in the control diet arm of a recent trial by Hall *et al*. (27). Discrepancies in energy balance are unsurprising, considering the cumulative error that would arise in comparisons encompassing three imprecise measures (energy intake, energy expenditure, and body energy stores), even with optimal conditions.

To elucidate underlying mechanisms involving diet and chronic disease, we will need a variety of complemental study designs, novel methods for ensuring high levels of dietary control for longer periods, multiple methods for measuring energy expenditure and substrate metabolism, and attention to effect modification by biological predisposition (2, 23, 28). Although research into more powerful behavioral and environmental interventions is also warranted, these approaches will be most effective when informed by accurate knowledge of the metabolic effects of dietary composition.

## Data Availability

The protocol and full dataset will be available at Open Science Framework upon peer review publication (https://osf.io/rvbuy/).

## Abbreviations

BMI: body mass index
CIM: carbohydrate-insulin model
DLW: doubly-labeled water
DXA: dual-energy x-ray absorptiometry
EER: estimated energy requirement for weight maintenance
Insulin-30: insulin concentration at 30 minutes following a 75-gram oral glucose load
ITT: Intention-to-Treat
PP: Per Protocol
TEE: total energy expenditure

## Acknowledgments

We thank Steven Heymsfield and Henry Feldman for critical feedback on the manuscript, and Stephanie Dickinson for verifying the data analyses. The study was funded by Nutrition Science Initiative (made possible by gifts from the Laura and John Arnold Foundation and Robert Lloyd Corkin Charitable Foundation), New Balance Foundation, Many Voices Foundation, and Blue Cross Blue Shield. DSL was supported by a mid-career mentoring award from the National Institute of Diabetes and Digestive and Kidney Diseases (K24DK082730).

## Author Contributions

CBE designed study, secured funding, interpreted data, participated in drafting the manuscript; LB helped design the study and analyzed the dietary records; PRL conducted the statistical analysis, interpreted data and participated in manuscript revision; GLK directed the parent and participated in manuscript revision; JMWW co-directed the parent study, helped design the Test diets and participated in manuscript revision; PKL co-directed the parent study at the performance site and participated in manuscript revision; WWW advised on double labeled water methodology and participated in manuscript revision; DSL, designed study, secured funding, interpreted data, participated in drafting the manuscript.

## Data Sharing

The protocol and full dataset is available at Open Science Framework (https://osf.io/rvbuy/).

## Supplementary Online Content

### TABLE OF CONTENTS

**Supplemental Table 1.** Changes in body composition by DXA and isotope dilution throughout the study. No significant diet effects were observed during the Test phase. Change in percentage fat by DXA was assessed as a difference, END – START. Change in percentage body fat by isotope dilution was assessed as a difference, average (MID, END) – START. Statistical models were minimally adjusted for cohort; adjustment for other baseline covariates did not materially affect the results.

| Intention-to-treat |  |  |  |  |  |  |  |
| --- | --- | --- | --- | --- | --- | --- | --- |
| Diet Group | N | Timepoint |  |  |  | Overall P | LO – HI<br>95% CI<br>P |
|  |  | PRE | START | MID | END |  |  |
| Percentage Body Fat from DXA |  |  |  |  |  |  |  |
| HIGH | 46 | 41.58 | 37.87 | – | 37.36 | 0.72 | 0.24 |
| MOD | 48 | 40.88 | 37.00 | – | 36.70 |  | -0.35 to 0.84 |
| LOW | 54 | 39.77 | 35.96 | – | 35.71 |  | 0.42 |
| Percentage Body Fat from isotope dilution |  |  |  |  |  |  |  |
| HIGH | 46 | 42.15 | 37.75 | 36.82 | 36.92 | 0.76 | -0.30 |
| MOD | 48 | 41.19 | 37.49 | 36.77 | 37.03 |  | -1.5 to 0.95 |
| LOW | 54 | 39.56 | 36.05 | 34.93 | 35.98 |  | 0.64 |
| Per protocol |  |  |  |  |  |  |  |
| Diet Group | N | Timepoint |  |  |  | Overall P | LO – HI<br>95% CI<br>P |
|  |  | PRE | START | MID | END |  |  |
| Percentage Body Fat from DXA |  |  |  |  |  |  |  |
| HIGH | 31 | 42.23 | 38.53 | – | 38.35 | 0.97 | -0.048 |
| MOD | 37 | 40.26 | 36.43 | – | 36.30 |  | -0.58 to 0.48 |
| LOW | 42 | 39.74 | 35.93 | – | 35.71 |  | 0.86 |
| Percentage Body Fat from isotope dilution |  |  |  |  |  |  |  |
| HIGH | 31 | 42.28 | 37.93 | 37.55 | 38.23 | 0.94 | -0.62 |
| MOD | 37 | 40.91 | 36.54 | 36.17 | 36.71 |  | -2.1 to 0.89 |
| LOW | 42 | 39.00 | 35.85 | 34.95 | 35.79 |  | 0.42 |

**Supplemental Figure 1.**
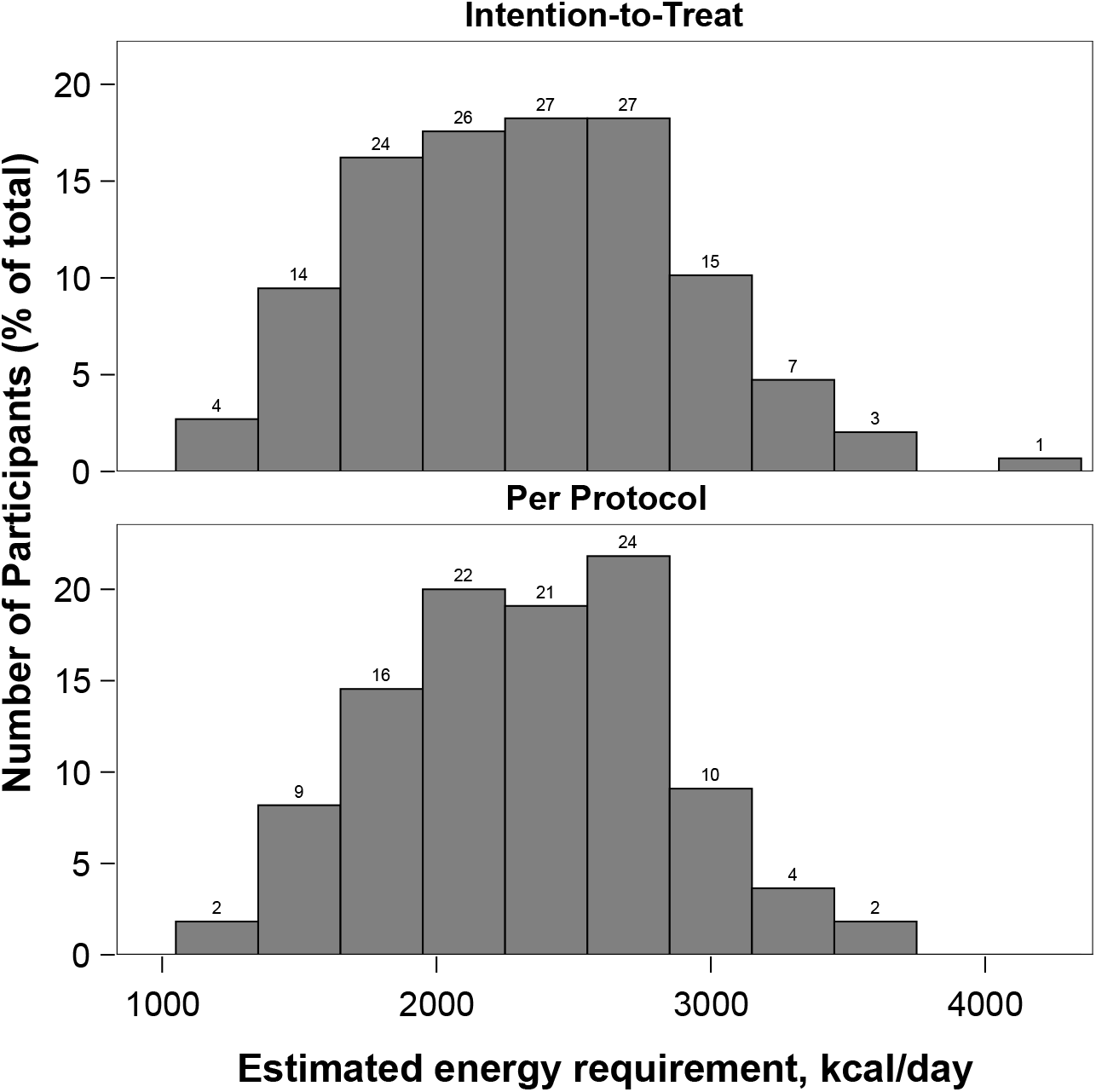
Distribution of estimated energy requirement (EER) in the Intention-to-Treat and Per Protocol analyses.

**Supplemental Figure 2.**
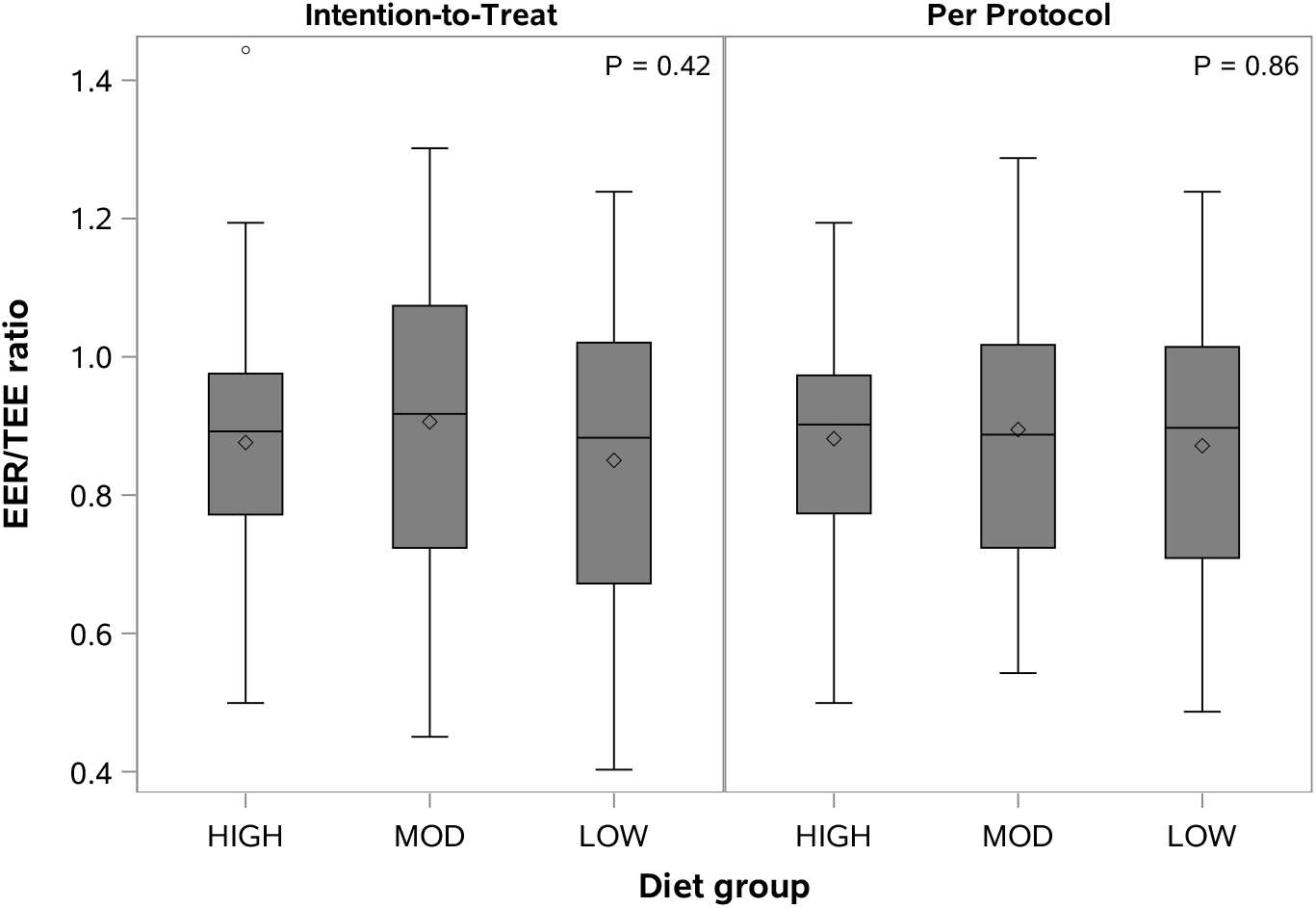
Ratio of estimated energy requirement (EER)-to-total energy expenditure (TEE) as a measure of non-adherence. Differences by diet group were not significant, suggesting no systematic bias. EER as a proportion of TEE in HIGH, MOD and LOW were, respectively: 0.88, 0.91, and 0.85 in the Intention-to-treat; and 0.88, 0.89 and 0.87 in the Per Protocol analyses. Symbols: diamonds, mean: horizontal lines, median; grey shaded area, interquartile range (25^th^ to 75^th^ percentile); bars, range (minimum to maximum).

